# Physician reimbursement and retention in HIV care: Racial disparities in the US South

**DOI:** 10.1101/2021.08.16.21262053

**Authors:** Zhongzhe Pan, Bassam Dahman, Rose S. Bono, Lindsay M. Sabik, Faye Z. Belgrave, Daniel E. Nixon, April D. Kimmel

**Affiliations:** Department of Health Behavior and Policy, Virginia Commonwealth University, Richmond, Virginia, USA; Department of Health Policy and Management, University of Pittsburgh Graduate School of Public Health, Pittsburgh, Pennsylvania, USA; Department of Psychology, Virginia Commonwealth University, Richmond, Virginia, USA; Department of Internal Medicine, Division of Infectious Diseases, Virginia Commonwealth University, Richmond, Virginia, USA

**Author notes:** **Corresponding author** April D. Kimmel, PhD, 830 East Main Street, 4th floor, Richmond, VA 23298.

## Abstract

Fewer than 60% of Americans diagnosed with HIV are retained in care, with racial disparities. Addressing structural barriers to care may improve outcomes along the HIV care continuum, such as retention, and promote health equity. We examined the relationship between physician reimbursement and retention in HIV care, including racial differences. Data included person-level demographic information and administrative claims (Medicaid Analytic eXtract, 2008-12), state Medicaid-to-Medicare fee ratios (Urban Institute, 2008, 2012), and county characteristics for 15 Southern states plus District of Columbia. The fee ratio is a standardized measure of physician reimbursement capturing state variation in Medicaid relative to Medicare physician reimbursement, which is largely consistent across states. We used generalized estimating equations to assess the association between physician reimbursement ratio and retention in HIV care (≥2 claims for physician visits, antiretroviral prescriptions, or CD4 or HIV RNA viral load tests ≥90 days apart in a calendar-year). We also evaluated an increase in the fee ratio to parity, where Medicaid and Medicare physician reimbursement are equal. Stratified analysis assessed racial differences. The sample included 55,237 adult Medicaid enrollees living with HIV (179,002 enrollee-years). Enrollees were retained in HIV care for approximately three-quarters (76.8%) of their enrollment-years, with retention lower among non-Hispanic Black (76.2%) versus non-Hispanic White (81.3%, *p*<0.001) enrollees. A 10-percentage point increase in physician reimbursement was associated with a 4% increase in the odds of retention (aOR 1.04, 95% CI 1.01, 1.08). In stratified analysis, increased physician reimbursement was significantly associated with retention among non-Hispanic Black but not non-Hispanic White enrollees. At parity, predicted retention was 81.1% (80.0%, 82.1%) and 80.4% (79%, 81.7%) of enrollment-years, overall and for non-Hispanic Black enrollees, respectively. Higher physician reimbursement improves retention in HIV care, particularly among non-Hispanic Black individuals living with HIV, and could be a structural mechanism to promote racial equity in retention.

## Introduction

Fewer than 60% of the estimated one million Americans diagnosed with HIV are retained in care,^1^ i.e., regular engagement with HIV-related medical care after diagnosis. Significant racial disparities exist, with non-Hispanic Blacks less likely to be retained in HIV care compared to non-Hispanic Whites.^2^ These differences may result in increased morbidity and mortality,^3^ decreased viral suppression,^4^ greater onward HIV transmission,^5^ and persistent racial inequity in access to care, quality of care, and health outcomes.^6^

Patient and provider interventions to increase retention in HIV care are well-studied,^7, 8^ but less attention has focused on how changes in structural-level factors influence retention or potential disparities. Structural factors are the economic, social, political, and institutional barriers or facilitators for health care access, delivery, quality of care, and health outcomes.^9^ Structural inequities disproportionately impact the health of racial and ethnic groups.^10^

Physician reimbursement, or payment from an insurer to a clinical provider for the provision of health services, is a structural factor that may impact quality of care and health outcomes. Lower physician reimbursement is associated with worse appointment availability,^11, 12^ longer waiting time,^13^ and lower probability of receiving care.^14^ Physician reimbursement has not been examined for HIV care quality, including retention.

We examined the association between physician reimbursement and retention in HIV care, and whether racial differences affected this relationship.

## Methods

### Data

We used the Medicaid Analytic eXtract (MAX), 2008-2012, for 15 Southern states plus District of Columbia.^15^ The MAX contains person-level administrative claims with information on eligibility, enrollment, demographics, outpatient visits (managed care encounters and fee-for-service), and filled prescriptions. The data comprise state-years meeting data quality and completeness metrics, which capture variability in encounter data reporting over time.^16^ We included county-level data on HIV prevalence (AIDSVu, 2015),^17^ urbanicity,^18^ and other county characteristics reflecting socioeconomic status and health care delivery (Area Health Resource Files, 2010).^19^ State-level Medicaid physician reimbursement was from Medicaid-to-Medicare fee ratio indices, 2008 and 2012.^20^

These data are among the best available to assess our research question. Medicaid is the largest single source of insurance for people living with HIV nationally.^1^ Additionally, Medicaid physician reimbursement is set at the state level, with substantial state variation^20^ and availability of reimbursement rates to researchers. This contrasts with other health insurers, such as Medicare (where physician reimbursement information is publicly available but rates are set centrally) and private insurers (where reimbursement data are proprietary). Further, the calendar-years used are contemporaneous due to lag times in release of Medicaid data, particularly in the more recent past as the Centers for Medicare and Medicaid Services transformed their Medicaid data collection system. Finally, the MAX have a similar demographic composition and basis of eligibility (income, disability) as current enrollees, including those living with HIV,^21^ with approximately 70% of enrollee and enrollee-years representing states that have not experienced Medicaid expansion.^22^ The MAX data used in the this analysis also reflect current medical practices regarding retention in care^23^ and historical definitions of the outcome (see below).^24, 25^

### Study population

The study population included Medicaid enrollees aged 19–64 years identified as living with HIV, eligible for Medicaid based on income or disability, and enrolled in Medicaid within a single state over the observation period (**Figure 1**). We used a case identification algorithm to identify enrollees living with HIV.^26^ To ensure complete utilization records, we excluded enrollees in a calendar-year, who were ever dually eligible for Medicare, ever enrolled in private insurance, not continuously enrolled throughout the calendar-year, ≥65 years, or had missing gender or county characteristics. We did not include Tennessee enrollees, since Tennessee’s Medicaid program did not have a fee-for-service component and therefore no information on physician reimbursement was available.^20^

**Figure 1.**
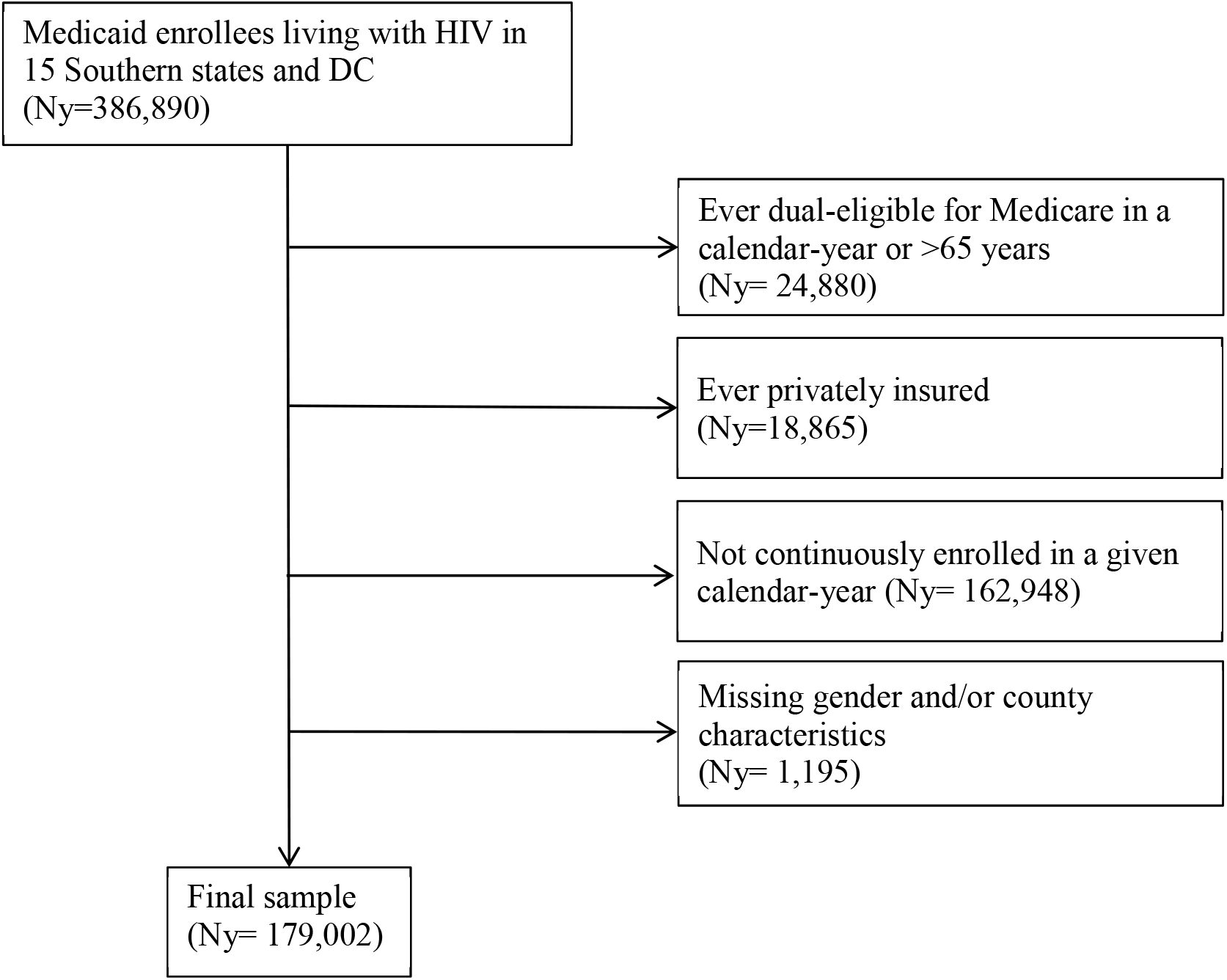
Sample selection process Medicaid enrollees living with HIV in 15 Southern states (Alabama, Arkansas, Delaware, Florida, Georgia, Kentucky, Lousiana, Maryland, Mississippi, North Carolina, Oklahoma, South Carolina, Texas, Virginia, West Virginia) and the District of Columbia were identified using an HIV identification algorithm.^26^ Enrollees were excluded from the sample if they were ever dually eligible for Medicare, older than 65 years, or ever privately insured, or with missing gender and/or county characteristics.

### Key variables

#### Retention in HIV care

The outcome variable was retention in HIV care^25^ and measured by calendar-year, January 1–December 31.^27^ The baseline measure (*Any Care Marker*) was defined as two or more care markers within a calendar-year, ≥90 days apart. Care markers included claims for routine HIV medical visits (i.e., outpatient office visit with HIV as the primary diagnosis), antiretroviral prescriptions, or HIV-related laboratory tests (e.g., CD4 count test). Care markers were identified in the claims using HIV diagnosis codes, service codes, and antiretroviral prescription drug codes. In the current study, 76.8% of total enrollee-years were retained in HIV care, slightly exceeding recent estimates.^27^

#### State-level physician reimbursement

Medicaid physician reimbursement was measured using the primary care Medicaid-to-Medicare fee ratio. This ratio compares physician reimbursement in the fee-for-service component of Medicaid with Medicare reimbursement for the same primary care services, by state.^20^ The ratios create a state-level index of relative Medicaid reimbursement levels, since Medicare physician reimbursement is typically higher than Medicaid physician reimbursement and intended to reflect costs of care, with limited geographic variation across states.^28^ Use of the Medicaid-to-Medicare fee ratio as a proxy for Medicaid physician reimbursement is consistent with the literature.^26, 29^ Medicaid-to-Medicare fee ratios were available for 2008 and 2012; we assumed a linear trend, applying estimated fee ratios for 2009, 2010, and 2011.^30^

#### Control variables

At the individual level, we included age at last observation in the observation period (grouped in 10-year intervals), gender, and race and ethnicity.^31^ We controlled for enrollee health status, including presence of co-infections (hepatitis B and/or C), non-AIDS comorbidities using the Charlson Comorbidity Index,^32^ and AIDS-defining illness.^33^ We also controlled for enrollee urbanicity, where counties in a federally designated metropolitan statistical area of ≥50,000 population were classified as urban and those in nonmetropolitan areas as rural.^18^ Additionally, we controlled for managed care enrollment, defined as enrollment ≥6 months in a calendar-year.^34^

At the county-level, we controlled for HIV prevalence, as well as physician availability and the number of hospital beds for non-specialized short-term care, which together were a proxy for availability of HIV clinical care.^35^ We additionally controlled for residential county-level socioeconomic status, including percent of the population with less than a high school diploma, percent currently unemployed, and median household income.^36^

### Statistical analysis

Our study is a based on longitudinal data where Medicaid enrollees were observed over years. We described sample characteristics overall and by race, testing racial differences using chi-squared tests for categorical variables and t-tests for continuous variables. When examining the relationship between physician reimbursement and retention in HIV care, we used generalized estimating equations considering potential within-person correlation of outcomes over multiple calendar-years of an individual’s Medicaid enrollment.^37, 38^ We specified a logit link, binomial distribution; the independent correlation matrix was found as the best fit model using the quasi information criterion. Given racial disparities in retention in HIV care,^39^ we performed stratified analysis for non-Hispanic White and non-Hispanic Black enrollees; we did not conduct analysis by Hispanic ethnicity due to small sample size and limited variation in the key variable of interest within this sub-sample. All models included state fixed effects to control for unobserved time-invariant heterogeneity in demographic or healthcare system characteristics across states, and year fixed effects to control for trends in retention in HIV care over time.

### Additional analysis

We applied alternative definitions of retention in care. We restricted care markers to CD4 cell count or HIV RNA viral load tests only (*Lab Tests*),^1^ as well as routine HIV medical care visits only (*Routine Visits*).^25^ Additionally, we considered an extended observation period of 15 months (i.e., January 1 of a given year through March 31 of the subsequent year) to account for real-world delays in medical visits and other services; here, we restricted the sample to those continuously enrolled in Medicaid for at least two years.

We considered separate sub-populations. We restricted the sample to enrollees living with HIV and requiring more intensive HIV management (*Intensive Management*). This sub-sample included enrollees who were pregnant, had HIV-related nephropathy, had an AIDS-defining illness, or received sulfamethoxazole/trimethoprim for *pneumocystis jiroveci* pneumonia treatment within a calendar-year.^40^ We also evaluated the relationship between physician reimbursement and retention in HIV care separately by enrollee payment type (i.e., fee-for-service versus managed care). This examined potential estimation biases associated with different service utilization patterns in the managed care versus fee-for-service components of Medicaid.

We also applied an alternative physician reimbursement index—the Medicaid-to-Medicare fee ratio for all services. This captured potential differences in practice patterns among specialist providers, including HIV clinicians. Finally, we examined the relationship at parity—i.e., a physician fee ratio of 1, indicating Medicaid and Medicare physician reimbursement are equal. We predicted the percentage retained in HIV care and change in the probability of retention, overall and by race, and holding covariates constant at their mean values.

Analyses were conducted in SAS version 9.4 and used two-sided tests with a significance threshold of *p*<0.05.

## Results

### Descriptive statistics

The sample included 55,273 enrollees living with HIV and 179,002 enrollee-years (**Table 1**). Enrollees were retained in care for over three-quarters of their total enrollment-years, with non-Hispanic White retained more than non-Hispanic Black enrollees (*p*<0.001). Compared to non-Hispanic Black enrollees, non-Hispanic White enrollees were older, more likely to be male, and had more co-infections, comorbidities, and AIDS-defining illnesses (*p*<0.01).

**Table 1.**
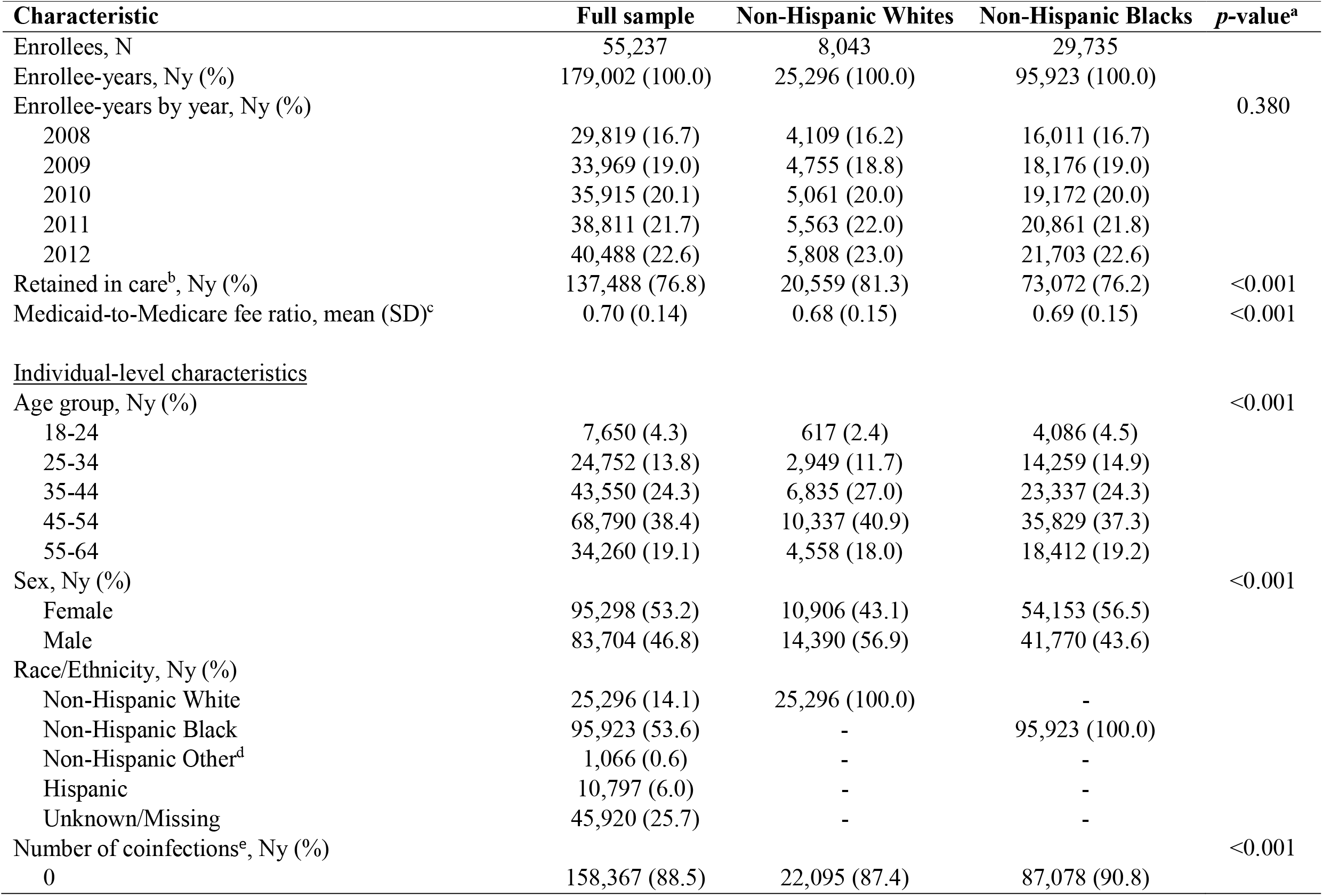
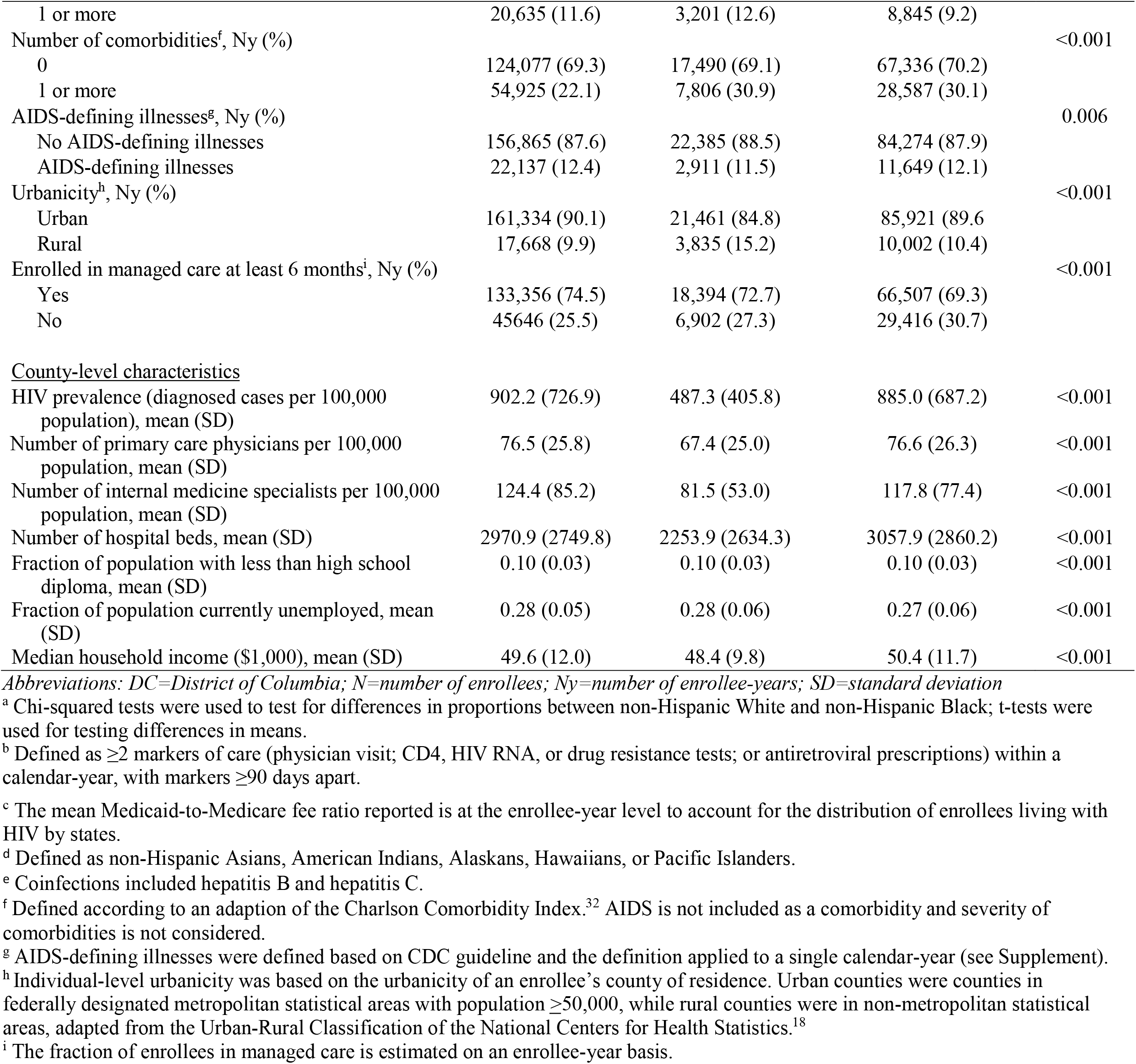
Descriptive statistics for Medicaid enrollees living with HIV in 15 Southern states plus DC, 2008-2012^a^

| Characteristic | Full sample | Non-Hispanic Whites | Non-Hispanic Blacks | p-value<sup>a</sup> |
| --- | --- | --- | --- | --- |
| Enrollees, N | 55,237 | 8,043 | 29,735 |  |
| Enrollee-years, Ny (%) | 179,002 (100.0) | 25,296 (100.0) | 95,923 (100.0) |  |
| Enrollee-years by year, Ny (%) |  |  |  | 0.380 |
| 2008 | 29,819 (16.7) | 4,109 (16.2) | 16,011 (16.7) |  |
| 2009 | 33,969 (19.0) | 4,755 (18.8) | 18,176 (19.0) |  |
| 2010 | 35,915 (20.1) | 5,061 (20.0) | 19,172 (20.0) |  |
| 2011 | 38,811 (21.7) | 5,563 (22.0) | 20,861 (21.8) |  |
| 2012 | 40,488 (22.6) | 5,808 (23.0) | 21,703 (22.6) |  |
| Retained in care<sup>b</sup>, Ny (%) | 137,488 (76.8) | 20,559 (81.3) | 73,072 (76.2) | <0.001 |
| Medicaid-to-Medicare fee ratio, mean (SD)<sup>c</sup> | 0.70 (0.14) | 0.68 (0.15) | 0.69 (0.15) | <0.001 |
| <u>Individual-level characteristics</u> |  |  |  |  |
| Age group, Ny (%) |  |  |  | <0.001 |
| 18-24 | 7,650 (4.3) | 617 (2.4) | 4,086 (4.5) |  |
| 25-34 | 24,752 (13.8) | 2,949 (11.7) | 14,259 (14.9) |  |
| 35-44 | 43,550 (24.3) | 6,835 (27.0) | 23,337 (24.3) |  |
| 45-54 | 68,790 (38.4) | 10,337 (40.9) | 35,829 (37.3) |  |
| 55-64 | 34,260 (19.1) | 4,558 (18.0) | 18,412 (19.2) |  |
| Sex, Ny (%) |  |  |  | <0.001 |
| Female | 95,298 (53.2) | 10,906 (43.1) | 54,153 (56.5) |  |
| Male | 83,704 (46.8) | 14,390 (56.9) | 41,770 (43.6) |  |
| Race/Ethnicity, Ny (%) |  |  |  |  |
| Non-Hispanic White | 25,296 (14.1) | 25,296 (100.0) | - |  |
| Non-Hispanic Black | 95,923 (53.6) | - | 95,923 (100.0) |  |
| Non-Hispanic Other<sup>d</sup> | 1,066 (0.6) | - | - |  |
| Hispanic | 10,797 (6.0) | - | - |  |
| Unknown/Missing | 45,920 (25.7) | - | - |  |
| Number of coinfections<sup>e</sup>, Ny (%) |  |  |  | <0.001 |
| 0 | 158,367 (88.5) | 22,095 (87.4) | 87,078 (90.8) |  |
| 1 or more | 20,635 (11.6) | 3,201 (12.6) | 8,845 (9.2) |  |
| Number of comorbidities<sup>f</sup>, Ny (%) |  |  |  | <0.001 |
| 0 | 124,077 (69.3) | 17,490 (69.1) | 67,336 (70.2) |  |
| 1 or more | 54,925 (22.1) | 7,806 (30.9) | 28,587 (30.1) |  |
| AIDS-defining illnesses<sup>g</sup>, Ny (%) |  |  |  | 0.006 |
| No AIDS-defining illnesses | 156,865 (87.6) | 22,385 (88.5) | 84,274 (87.9) |  |
| AIDS-defining illnesses | 22,137 (12.4) | 2,911 (11.5) | 11,649 (12.1) |  |
| Urbanicity<sup>h</sup>, Ny (%) |  |  |  | <0.001 |
| Urban | 161,334 (90.1) | 21,461 (84.8) | 85,921 (89.6) |  |
| Rural | 17,668 (9.9) | 3,835 (15.2) | 10,002 (10.4) |  |
| Enrolled in managed care at least 6 months<sup>i</sup>, Ny (%) |  |  |  | <0.001 |
| Yes | 133,356 (74.5) | 18,394 (72.7) | 66,507 (69.3) |  |
| No | 45,646 (25.5) | 6,902 (27.3) | 29,416 (30.7) |  |
| <u>County-level characteristics</u> |  |  |  |  |
| HIV prevalence (diagnosed cases per 100,000 population), mean (SD) | 902.2 (726.9) | 487.3 (405.8) | 885.0 (687.2) | <0.001 |
| Number of primary care physicians per 100,000 population, mean (SD) | 76.5 (25.8) | 67.4 (25.0) | 76.6 (26.3) | <0.001 |
| Number of internal medicine specialists per 100,000 population, mean (SD) | 124.4 (85.2) | 81.5 (53.0) | 117.8 (77.4) | <0.001 |
| Number of hospital beds, mean (SD) | 2970.9 (2749.8) | 2253.9 (2634.3) | 3057.9 (2860.2) | <0.001 |
| Fraction of population with less than high school diploma, mean (SD) | 0.10 (0.03) | 0.10 (0.03) | 0.10 (0.03) | <0.001 |
| Fraction of population currently unemployed, mean (SD) | 0.28 (0.05) | 0.28 (0.06) | 0.27 (0.06) | <0.001 |
| Median household income (\$1,000), mean (SD) | 49.6 (12.0) | 48.4 (9.8) | 50.4 (11.7) | <0.001 |

*Abbreviations: DC=District of Columbia; N=number of enrollees; Ny=number of enrollee-years; SD=standard deviation*
<sup>a</sup> Chi-squared tests were used to test for differences in proportions between non-Hispanic White and non-Hispanic Black; t-tests were used for testing differences in means.
<sup>b</sup> Defined as $\geq 2$ markers of care (physician visit; CD4, HIV RNA, or drug resistance tests; or antiretroviral prescriptions) within a calendar-year, with markers $\geq 90$ days apart.
<sup>c</sup> The mean Medicaid-to-Medicare fee ratio reported is at the enrollee-year level to account for the distribution of enrollees living with HIV by states.
<sup>d</sup> Defined as non-Hispanic Asians, American Indians, Alaskans, Hawaiians, or Pacific Islanders.
<sup>e</sup> Coinfections included hepatitis B and hepatitis C.
<sup>f</sup> Defined according to an adaption of the Charlson Comorbidity Index.<sup>32</sup> AIDS is not included as a comorbidity and severity of comorbidities is not considered.
<sup>g</sup> AIDS-defining illnesses were defined based on CDC guideline and the definition applied to a single calendar-year (see Supplement).
<sup>h</sup> Individual-level urbanicity was based on the urbanicity of an enrollee's county of residence. Urban counties were counties in federally designated metropolitan statistical areas with population $\geq 50,000$ , while rural counties were in non-metropolitan statistical areas, adapted from the Urban-Rural Classification of the National Centers for Health Statistics.<sup>18</sup>
<sup>i</sup> The fraction of enrollees in managed care is estimated on an enrollee-year basis.

### Main analysis

We found a positive and significant relationship between physician reimbursement and retention in HIV care (**Table 2**). A 10-percentage point increase in the Medicaid-to-Medicare fee ratio was associated with a 4% increase in the likelihood of retention in care (adjusted odds ratio 1.04; 95% confidence interval 1.01, 1.08). In stratified analysis, there was an association only among non-Hispanic Black enrollees (1.08; 1.05, 1.12).

**Table 2.**
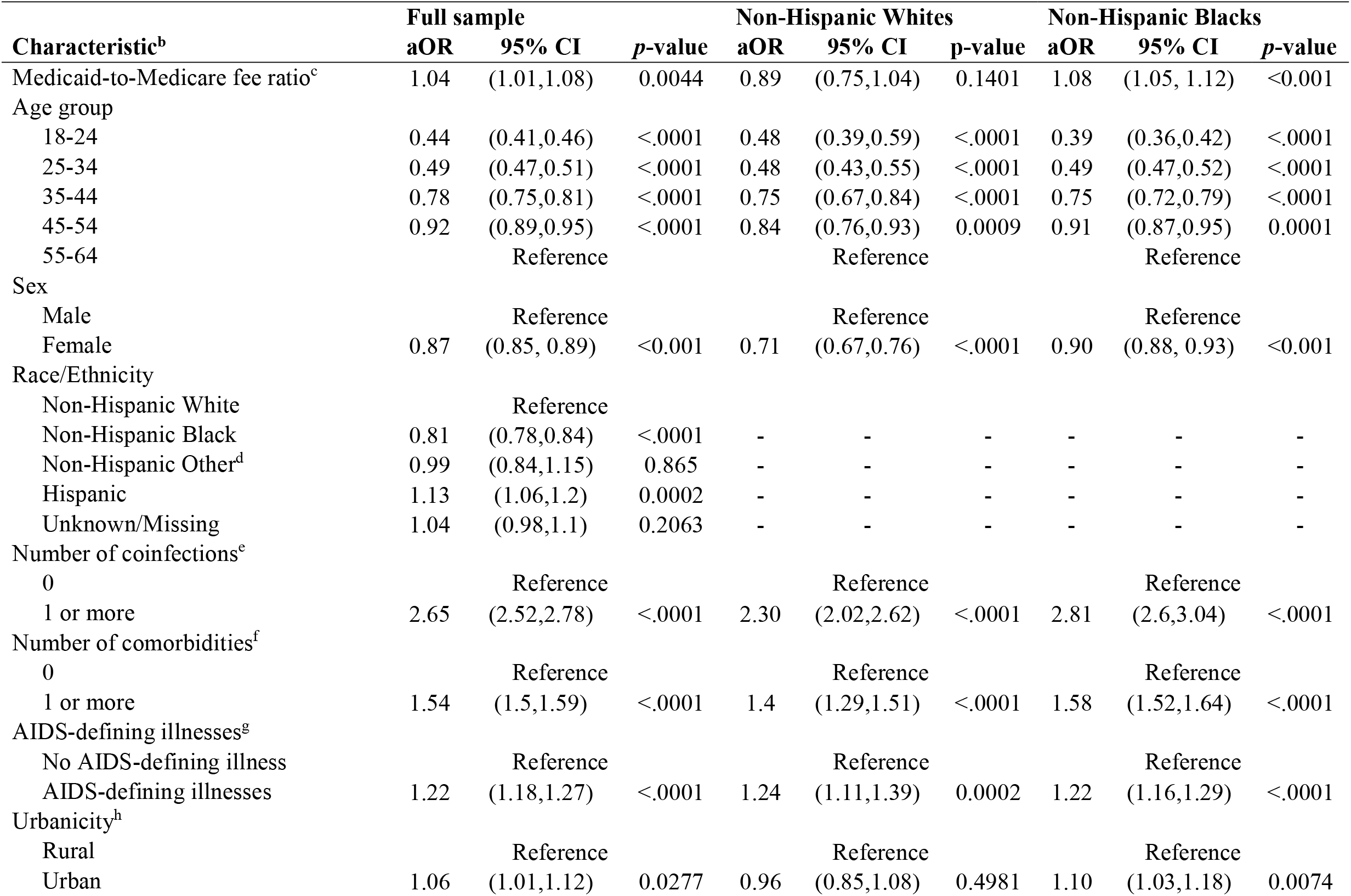
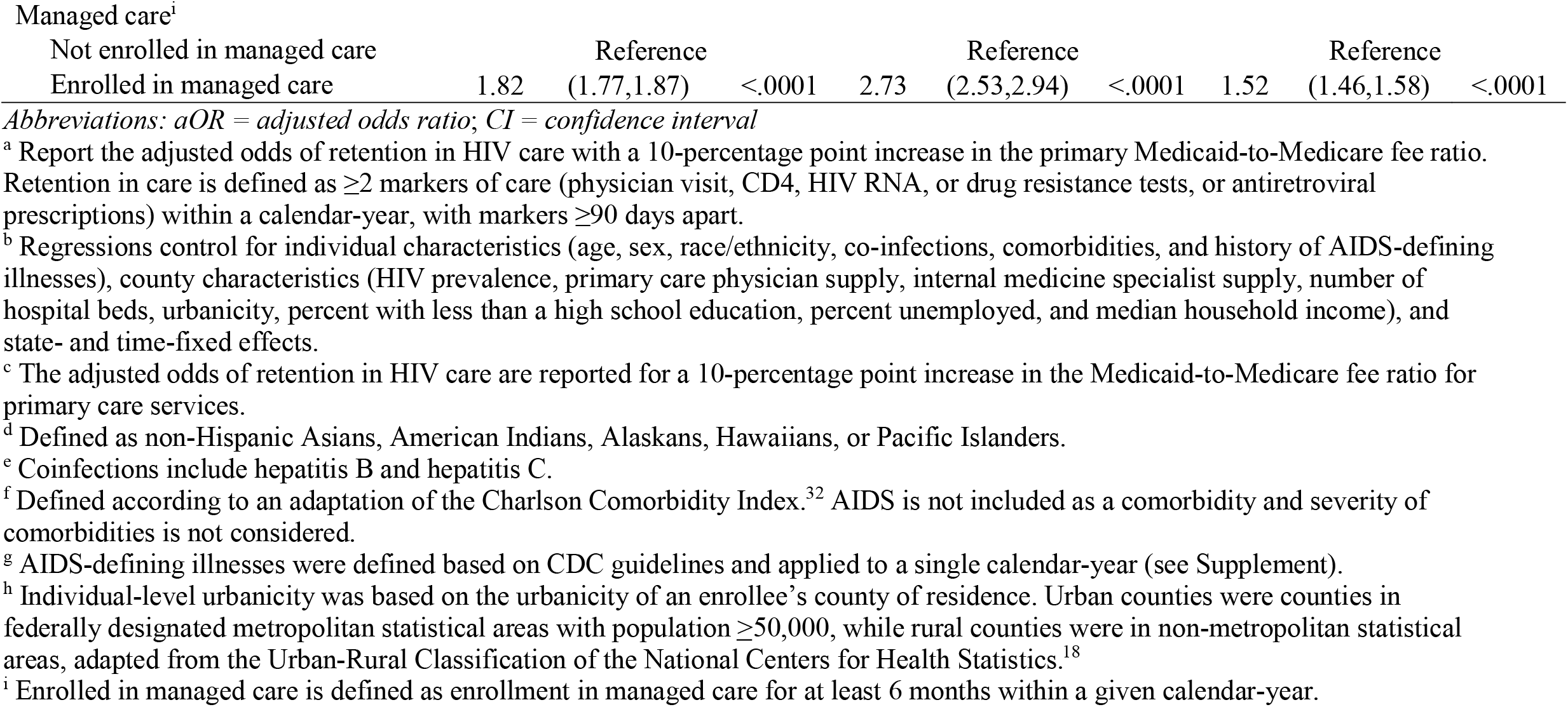
State-level Medicaid physician reimbursement and retention in HIV care: baseline analysis^a^

| **Characteristic<sup>b</sup>** | **Full sample** |  |  | **Non-Hispanic Whites** |  |  | **Non-Hispanic Blacks** |  |  |
| --- | --- | --- | --- | --- | --- | --- | --- | --- | --- |
|  | **aOR** | **95% CI** | **p-value** | **aOR** | **95% CI** | **p-value** | **aOR** | **95% CI** | **p-value** |
| Medicaid-to-Medicare fee ratio<sup>c</sup> | 1.04 | (1.01,1.08) | 0.0044 | 0.89 | (0.75,1.04) | 0.1401 | 1.08 | (1.05, 1.12) | <0.001 |
| Age group |  |  |  |  |  |  |  |  |  |
| 18-24 | 0.44 | (0.41,0.46) | <.0001 | 0.48 | (0.39,0.59) | <.0001 | 0.39 | (0.36,0.42) | <.0001 |
| 25-34 | 0.49 | (0.47,0.51) | <.0001 | 0.48 | (0.43,0.55) | <.0001 | 0.49 | (0.47,0.52) | <.0001 |
| 35-44 | 0.78 | (0.75,0.81) | <.0001 | 0.75 | (0.67,0.84) | <.0001 | 0.75 | (0.72,0.79) | <.0001 |
| 45-54 | 0.92 | (0.89,0.95) | <.0001 | 0.84 | (0.76,0.93) | 0.0009 | 0.91 | (0.87,0.95) | 0.0001 |
| 55-64 |  | Reference |  |  | Reference |  |  | Reference |  |
| Sex |  |  |  |  |  |  |  |  |  |
| Male |  | Reference |  |  | Reference |  |  | Reference |  |
| Female | 0.87 | (0.85, 0.89) | <0.001 | 0.71 | (0.67,0.76) | <.0001 | 0.90 | (0.88, 0.93) | <0.001 |
| Race/Ethnicity |  |  |  |  |  |  |  |  |  |
| Non-Hispanic White |  | Reference |  |  |  |  |  |  |  |
| Non-Hispanic Black | 0.81 | (0.78,0.84) | <.0001 | - | - | - | - | - | - |
| Non-Hispanic Other<sup>d</sup> | 0.99 | (0.84,1.15) | 0.865 | - | - | - | - | - | - |
| Hispanic | 1.13 | (1.06,1.2) | 0.0002 | - | - | - | - | - | - |
| Unknown/Missing | 1.04 | (0.98,1.1) | 0.2063 | - | - | - | - | - | - |
| Number of coinfections<sup>e</sup> |  |  |  |  |  |  |  |  |  |
| 0 |  | Reference |  |  | Reference |  |  | Reference |  |
| 1 or more | 2.65 | (2.52,2.78) | <.0001 | 2.30 | (2.02,2.62) | <.0001 | 2.81 | (2.6,3.04) | <.0001 |
| Number of comorbidities<sup>f</sup> |  |  |  |  |  |  |  |  |  |
| 0 |  | Reference |  |  | Reference |  |  | Reference |  |
| 1 or more | 1.54 | (1.5,1.59) | <.0001 | 1.4 | (1.29,1.51) | <.0001 | 1.58 | (1.52,1.64) | <.0001 |
| AIDS-defining illnesses<sup>g</sup> |  |  |  |  |  |  |  |  |  |
| No AIDS-defining illness |  | Reference |  |  | Reference |  |  | Reference |  |
| AIDS-defining illnesses | 1.22 | (1.18,1.27) | <.0001 | 1.24 | (1.11,1.39) | 0.0002 | 1.22 | (1.16,1.29) | <.0001 |
| Urbanicity<sup>h</sup> |  |  |  |  |  |  |  |  |  |
| Rural |  | Reference |  |  | Reference |  |  | Reference |  |
| Urban | 1.06 | (1.01,1.12) | 0.0277 | 0.96 | (0.85,1.08) | 0.4981 | 1.10 | (1.03,1.18) | 0.0074 |
| Managed care<sup>i</sup> |  |  |  |  |  |  |  |  |  |
| Not enrolled in managed care |  | Reference |  |  | Reference |  |  | Reference |  |
| Enrolled in managed care | 1.82 | (1.77,1.87) | <.0001 | 2.73 | (2.53,2.94) | <.0001 | 1.52 | (1.46,1.58) | <.0001 |

Abbreviations: aOR = adjusted odds ratio; CI = confidence interval
<sup>a</sup> Report the adjusted odds of retention in HIV care with a 10-percentage point increase in the primary Medicaid-to-Medicare fee ratio. Retention in care is defined as $\geq 2$ markers of care (physician visit, CD4, HIV RNA, or drug resistance tests, or antiretroviral prescriptions) within a calendar-year, with markers $\geq 90$ days apart.
<sup>b</sup> Regressions control for individual characteristics (age, sex, race/ethnicity, co-infections, comorbidities, and history of AIDS-defining illnesses), county characteristics (HIV prevalence, primary care physician supply, internal medicine specialist supply, number of hospital beds, urbanicity, percent with less than a high school education, percent unemployed, and median household income), and state- and time-fixed effects.
<sup>c</sup> The adjusted odds of retention in HIV care are reported for a 10-percentage point increase in the Medicaid-to-Medicare fee ratio for primary care services.
<sup>d</sup> Defined as non-Hispanic Asians, American Indians, Alaskans, Hawaiians, or Pacific Islanders.
<sup>e</sup> Coinfections include hepatitis B and hepatitis C.
<sup>f</sup> Defined according to an adaptation of the Charlson Comorbidity Index.<sup>32</sup> AIDS is not included as a comorbidity and severity of comorbidities is not considered.
<sup>g</sup> AIDS-defining illnesses were defined based on CDC guidelines and applied to a single calendar-year (see Supplement).
<sup>h</sup> Individual-level urbanicity was based on the urbanicity of an enrollee's county of residence. Urban counties were counties in federally designated metropolitan statistical areas with population $\geq 50,000$ , while rural counties were in non-metropolitan statistical areas, adapted from the Urban-Rural Classification of the National Centers for Health Statistics.<sup>18</sup>
<sup>i</sup> Enrolled in managed care is defined as enrollment in managed care for at least 6 months within a given calendar-year.

### Additional analysis

Baseline findings were robust to alternative definitions of retention in care and the physician reimbursement index (**Figure 2**). When using the *Lab Tests* definition of retention, a 10-percentage point increase in the fee ratio was associated with a 21% increase (1.18, 1.24) in the adjusted odds of retention in care. Similarly, when using the *Routine Visits* definition of retention, a 10-percentage point increase in the fee ratio was associated with a 10% increase (1.07, 1.13) in the adjusted odds of retention in care. The relationship remained significant when extending the observation period for retention to 15 months. In stratified analysis, the relationship was statistically significant among non-Hispanic Black enrollees, regardless of definition of retention in HIV care; similar to the stratified analysis in the main analysis, there was no statistically significant relationship among non-Hispanic White enrollees. When using the fee ratio for all services, versus primary care services only, the relationship for the full sample and stratified analysis was similar to the baseline findings.

**Figure 2.**
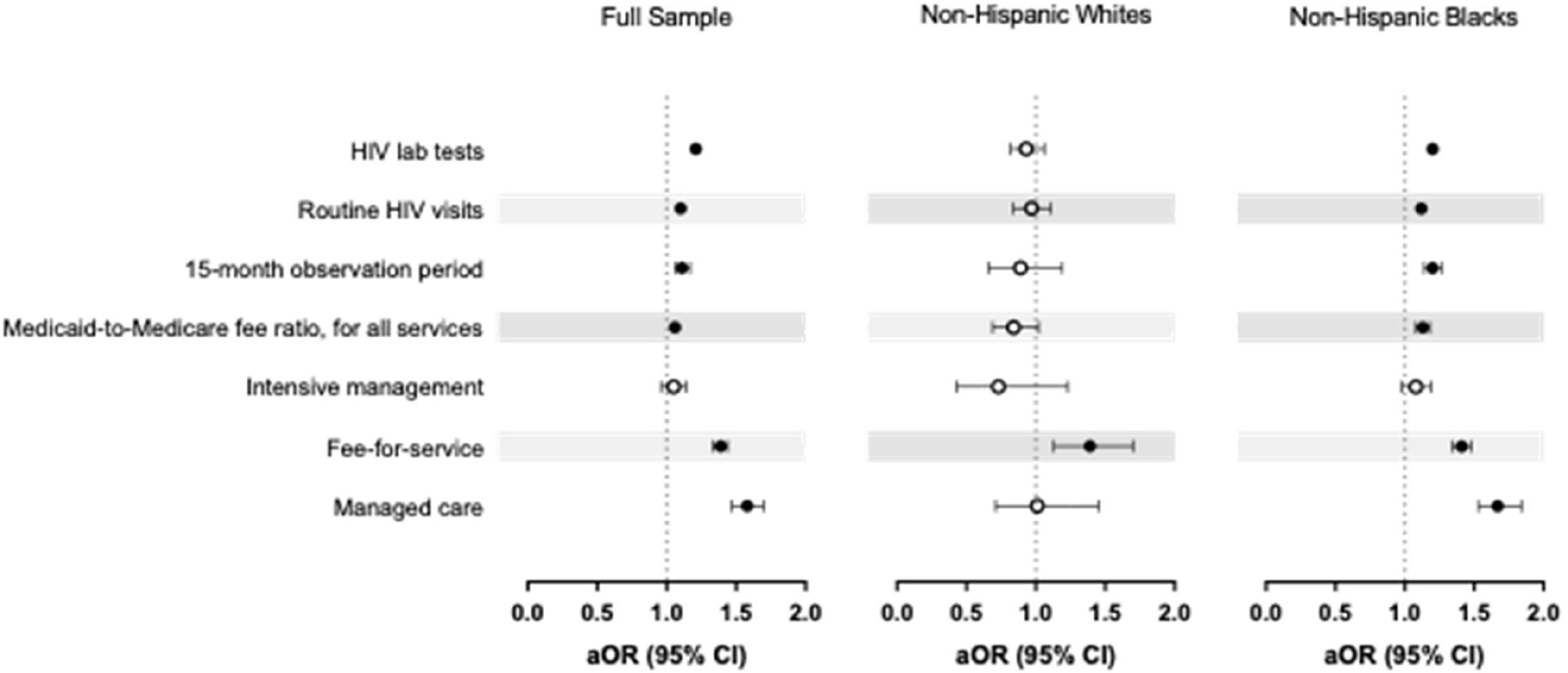
State-level Medicaid physician reimbursement and retention in HIV care: summary of sensitivity analyses *Abbreviations: aOR = adjusted odds ratio*; *CI=confidence interval* Reported are the adjusted odds of retention in HIV care with a 10-percentage point increase in the primary Medicaid-to-Medicare fee ratio. Retention in care is defined as ≥2 markers of care (physician visit, CD4, HIV RNA, or drug resistance tests, or antiretroviral prescriptions) within a calendar-year, with markers ≥90 days apart. Regressions control for individual characteristics (age, sex, race/ethnicity, co-infections, comorbidities, and history of AIDS-defining illnesses), county characteristics (HIV prevalence, primary care physician supply, internal medicine specialist supply, number of hospital beds, urbanicity, percent with less than a high school education, percent unemployed, and median household income), and state- and time-fixed effects. *HIV lab tests* refers to ≥2 CD4 or HIV RNA tests within a calendar-year, with markers ≥90 days apart. *Routine HIV visits* refers to ≥2 physician visit within a calendar-year, with markers ≥90 days apart. *15-month observation period* refers to ≥2 markers of care (physician visit, CD4, HIV RNA, or drug resistance tests, or antiretroviral prescriptions) within 15-month period (a calendar-year plus the first three months in the subsequent calendar-year), with markers ≥90 days apart. The sample used for this sensitivity analysis is restricted to enrollees continuously enrolled in Medicaid for two years. *Intensive management* refers to a sub-sample of enrollees who have evidence of requiring intensive HIV management (due to pregnancy, HIV-related nephropathy, an AIDS-defining illness, or receipt of sulfamethoxazole/trimethoprim for *pneumocystis jiroveci* pneumonia) in a given calendar-year. *Fee-for-service* refers to a sub-sample of enrollees who have no evidence of managed care enrollment in a given calendar-year. *Managed care* refers to a sub-sample of enrolled in comprehensive care for at least 6 months in a given calendar-year.

Findings were mixed when examining the relationship in different sub-populations. In the *Intensive Management* sample, we found no significant relationship between physician reimbursement and retention in care. However, for both the Medicaid fee-for-service and the Medicaid managed care sub-populations, a 10-percentage point increase in the fee ratio was positively and significantly associated with retention in care. In stratified analysis, findings were similar for non-Hispanic Black enrollees. We found no significant relationship among non-Hispanic White enrollees across all sensitivity analyses, with the exception of fee-for-service enrollees.

When predicting an increase to parity, retention in HIV care increased overall by 2.7% (0.6%, 4.7%) compared to the main analysis, with enrollees retained in care for 81.1% of enrollment-years. For non-Hispanic Black enrollees, retention increased by 5.4% (2.4%, 8.3%), with 80.4% of their enrollee-years retained in HIV care. Results were not significant for non-Hispanic White enrollees.

## Discussion

Retention in HIV care, and racial disparities in retention, remain ongoing US challenges that could be mitigated by addressing structural barriers to care. This study finds that retention in HIV care is higher among enrollees living in states with higher Medicaid physician reimbursement, regardless of different definitions of care retention, enrollee payment type, and type of fee ratio. Notably, in stratified analyses, this relationship occurs only among non-Hispanic Black Medicaid enrollees.

Our findings suggest that even modest increases in physician reimbursement may improve retention in HIV care. More sizeable increases have been implemented, but only in the relative short-term. For example, the Affordable Care Act increased state-level Medicaid physician reimbursement for primary care services on average 73% nationally during 2013-14 to promote primary care physician participation in Medicaid.^20^ Recently, 37 states have temporarily increased Medicaid provider reimbursement for certain providers in response to the COVID-19 public health emergency.^22, 41^ These temporary increases suggest that states recognize the importance of Medicaid reimbursement in supporting access to care, but may face constraints that preclude committing to substantial payment changes over the longer term. We consider a more modest 10-percentage point increase in state-level Medicaid physician reimbursement—which may be economically feasible for some state Medicaid programs. This increase was associated with a 4%–39% increase in the odds of retention in HIV care, with potential downstream improvements: increased retention in HIV care may improve HIV RNA viral suppression,^4^ thereby reducing morbidity, mortality,^3^ and HIV transmission.^5, 42^

Increasing physician reimbursement may reduce racial disparities in retention in HIV care and improve health equity. Increased physician reimbursement was associated with higher retention among non-Hispanic Black—but not non-Hispanic White—enrollees in stratified analyses. One explanation for racial differences in the association is the manifold structural disparities inherently faced by Blacks. Reduction in one structural barrier might have a disproportionately more favorable impact for non-Hispanic Black than non-Hispanic White individuals living with HIV, who face fewer structural barriers. The removal of one structural barrier is likely to have greater weight for groups burdened by many structural barriers; that is, our results may reflect a ceiling effect among non-Hispanic White enrollees, who had a higher proportion of their enrollment retained in care (81%) than non-Hispanic Black enrollees (76%). Indeed, increasing Medicaid physician reimbursement to parity may be a mechanism to narrow the gap between non-Hispanic Whites and non-Hispanic Blacks retained in HIV care.

Another explanation is that the providers in practices with larger shares of racial and ethnic minority patients may benefit more from increased physician reimbursement. Evidence suggests that practices with higher shares of racial and ethnic minority patients (>70%) are more likely to have patients participating in Medicaid, be located in states with lower Medicaid-to-Medicare fee ratios, and be located in communities with lower private insurance reimbursement.^43^ Providers in these practices have reported care challenges—including shorter visit length, delays in reporting from other providers, and a higher scope of care without specialty referrals—that influence quality of care, care coordination, and possibly care retention.^43^ Other research indicates that clinics serving at least 30% racial and ethnic minority patients have medically and psychosocially more complex patients, lower access to medical supplies and referral specialists, fewer exam rooms per physician, and a more chaotic work environment, with physicians reporting lower control of their work and job satisfaction.^44^ Lower physician reimbursement may therefore function as an unseen structural barrier to racial equity in health care.^45^

This study is the first to examine the relationship between physician reimbursement and HIV care quality, building on the broader literature on physician reimbursement as a potential system-level barrier to high quality care. This study’s findings among non-Hispanic Black Medicaid enrollees living with HIV align with previous work indicating that higher physician reimbursement is associated with more frequent physician visits,^14, 29^ increased usual source of care,^29^ and outpatient department visits.^14, 29^ Study findings also parallel the limited research on physician reimbursement for HIV care: over half of general internists reported that higher physician reimbursement for one clinical service (counseling time) would facilitate routine HIV screening,^46^ a key element of the HIV care continuum. Finally, our work is among the first to examine the differential impact of physician reimbursement on quality of care by race. Disentangling how upstream factors shape disparities in quality of care is critical to undermining structural racism and ending racial disparities in health and healthcare.

## Limitations

First, the fee ratio reflects physician reimbursement for Medicaid fee-for-service versus Medicaid managed care. However, physician reimbursement for managed care approximates fee-for-service reimbursement.^47^ Second, we conducted the analysis using the Medicaid-to-Medicare fee ratio for primary care services, which does not fully align with the care markers used to define retention in HIV care. This index also may not capture changes in utilization of specialist services, particularly among non-Hispanic White enrollees with HIV who are more likely to see infectious disease specialists for their usual care.^48^ However, our findings were robust when examining the Medicaid-to-Medicare fee ratio for all services, which captures specialist practice patterns. Third, our measure of physician reimbursement cannot capture payments clinicians may receive from the federal Ryan White HIV/AIDS Program for services provided to Medicaid enrollees but not reimbursed by Medicaid, although this is unlikely in our sample.

Fourth, a calendar-year definition of retention in care may underestimate enrollee retention.^25^ Administrative claims, versus outcomes from clinical trials or cohort studies, provide a real-world snapshot of health system interactions, including variation in appointment availability and delays in obtaining care. We account for this reality when extending the observation period for retention in HIV care from 12 to 15 months, with similar findings. Fifth, we could not control for variation in provider- or clinic-level characteristics, as claims data do not allow precise identification of an enrollee’s primary provider of record (including primary HIV provider). We also could not control for all relevant social and behavioral barriers to retention in care,^49^ since the data do not include information on individual social determinants of health (e.g., housing insecurity). Moreover, limitations in Medicaid coverage for substance use or mental health care prior to ACA implementation coupled with wide state-level variation in such coverage precluded accurate identification of substance use and behavioral health disorders.^50, 51^ Finally, we conducted the analysis based on a single geographic region, the US South. Since physician reimbursement in most Southern states is higher than the national average (see Supplement),^20^ we anticipate that our findings would generalize beyond the South.

## Conclusions

Increasing physician reimbursement may be a feasible policy solution to improve, and promote racial equity in, quality of care for people living with HIV. Addressing structural barriers such as low physician reimbursement may improve retention in HIV care among non-Hispanic Black individuals living with HIV, improve racial equity in retention, and facilitate achievement of national goals to improve health outcomes in this population.

## Data Availability

The Medicaid Analytic eXtract (MAX) data files are available from the Centers for Medicare and Medicaid Services via the Research Data Assistance Center (ResDAC).

## Acknowledgements

This work was supported in part by the National Institutes of Health (award number R01 MD11277) and the Blick Scholars Program at Virginia Commonwealth University. The funders did not contribute to the design and conduct of the study; collection, management, analysis, and interpretation of the data; preparation, review, or approval of the manuscript; and decision to submit the manuscript for publication. There are no known conflicts of interest, financial or otherwise. This work was previously presented in part at the AcademyHealth Annual Research Meeting (abstract number 439, July 28–August 6, 2020, Virtual meeting).

## Notes

### Competing Interest Statement

The authors have declared no competing interest.

### Author Declarations

The IRB that provided approval for the study was the Virginia Commonwealth University Institutional Review Board. Ethical approval was given for the research.

